# Epigenetic signatures of biological vulnerability and residual risk in heart failure

**DOI:** 10.64898/2026.09.01.26361996

**Authors:** Pascal B. Meyre, Michael Chong, Elad Shemesh, Shihong Mao, Sukrit Narula, Ambuj Roy, Kamilu M. Karaye, Georg Ertl, Lisa Mielniczuk, Sanjib Kumar Sharma, Bishav Mohan, Fernando Lanas, Thomas Wittlinger, Ahmet Celik, Jabir Abdullakutty, Julio Núñez, Okechukwu S. Ogah, Narendra Jathappa, Andrzej Budaj, Aldo P. Maggioni, Jean Rouleau, Kumar Balasubramanian, Tara McCready, Alex Grinvalds, Darryl P. Leong, Philip G. Joseph, Salim Yusuf, Guillaume Paré

**Affiliations:** Population Health Research Institute, McMaster University, Hamilton, ON, Canada; Cardiovascular Research Institute Basel and Department of Cardiology, University Hospital Basel, University of Basel, Switzerland; Department of Pathology and Molecular Medicine, McMaster University, Hamilton, Canada; Department of Internal Medicine, Yale University, New Haven, Connecticut, United States of America; Department of Cardiology, All India Institute of Medical Sciences, New Delhi, Delhi, India; Bayero University and Aminu Kano Teaching Hospital, Department of Medicine, Kano, Nigeria; Comprehensive Heart Failure Center, University Hospital, Würzburg, Germany; Mayo Clinic, Department of Cardiovascular Medicine, Rochester, MN, USA; BP Koirala Institute of Health Sciences, Dharan, Kathmandu, Nepal; Dayanand Medical College and Hospital, Cardiology, Ludhiana, India; Universidad de La Frontera, Temuco, Chile; Department of Cardiology, Asklepios Hospital Goslar, Goslar, Germany; Faculty of Medicine, Mersin University, Mersin, Türkiye; Lisie Hospital, Kerala, India; Servicio de Cardiología, Hospital Clínico Universitario Valencia, Valencia, Spain and fundación para la Investigación del Hospital Clínico de la Comunidad Valenciana (INCLIVA), Valencia, Spain; Cardiology Unit, Department of Medicine, University of Ibadan/University College Hospital, Ibadan, Oyo State, Nigeria; Nanjappa Life Care, Karnataka, India; Centre of Postgraduate Medical Education, Department of Cardiology, Grochowski Hospital, Warsaw, Poland; ANMCO Research Center, Heart Care Foundation, Florence, Italy; Montreal Heart Institute, Université de Montréal, Montreal, Quebec, Canada; Thrombosis and Atherosclerosis Research Institute, Hamilton Health Sciences and McMaster University, Hamilton, Canada

**Keywords:** Heart failure, DNA methylation, Epigenetics, Aging clocks, Biological aging, Mortality

## Abstract

**Background:** DNA methylation (DNAm) signatures capture cumulative lifestyle exposures and biological aging. This prospective study evaluated whether DNAm-based scores and epigenetic aging clocks are associated with clinical outcomes and mortality in a multinational cohort of patients with heart failure (HF).

**Methods:** We studied 2,594 patients with HF from 40 countries in the Global Congestive Heart Failure (G-CHF) registry with whole-blood DNAm data. Fifteen published DNAm-based scores and epigenetic aging clocks reflecting lifestyle, environmental and physiological exposures, inflammation, frailty, mortality risk, and biological aging were derived. Associations with HF hospitalization, cardiovascular death, and all-cause death were assessed using multivariable Cox regression adjusted for age, sex, ancestry, the MAGGIC risk score, and NT-proBNP. Incremental prognostic value was compared to MAGGIC score and NT-proBNP. Extreme DNAm profiles were defined as scores or clocks exceeding ±1.5 standard deviations (s.d.) from the population mean.

**Results:** Mean age was 62.7±14.0 years, 66.2% were male, and mean left ventricular ejection fraction was 40.1±14.1%. During a median follow-up of 3.0 years, 338 patients were hospitalized for HF, 349 died from cardiovascular causes, and 565 died from any cause. Higher epigenetic age and DNAm scores for CRP, frailty, and mortality were associated with increased risk, whereas higher diet-related DNAm scores were inversely associated. For all-cause death, adjusted hazard ratios per 1-s.d. were 1.36 (95% CI, 1.24-1.50) for GrimAge, 1.27 (95% CI, 1.17-1.38) for the DNAm score for CRP, 1.44 (95% CI, 1.28-1.63) for the DNAm score for frailty, and 1.48 (95% CI, 1.32-1.65) for the DNAm score for mortality, compared with 0.81 (95% CI, 0.75-0.88) and 0.86 (95% CI, 0.79-0.94) for the DNAm scores for Alternative Healthy Eating Index and Mediterranean Diet Score. These patterns were directionally consistent for cardiovascular death and weaker for HF hospitalization and were more pronounced among patients with lower clinical risk (MAGGIC<17, P_interaction_<0.05), particularly for all-cause death. Patients with 4-5 extreme-high DNAm scores or clocks had more than twice the risk of death (HR, 2.27; 95% CI, 1.65-3.13).

**Conclusions:** DNAm-based scores and epigenetic aging clocks reflect multiple dimensions of biological vulnerability in HF and are associated with clinical outcomes and mortality beyond clinical risk factors.

**Clinical Perspective:** *What is new?:*

- In 2,594 patients with heart failure (HF) from 40 countries, blood-based DNA methylation (DNAm) scores for inflammation, frailty, and mortality, as well as epigenetic aging measures, were associated with mortality and other clinical outcomes after adjustment for established risk factors and NT-proBNP.
- These associations were stronger among patients with lower clinical risk, suggesting that epigenetic signatures may capture biological vulnerability not fully reflected by conventional risk assessment.
- A greater accumulation of adverse epigenetic signatures was associated with progressively higher mortality risk; each additional extreme-high DNAm score or aging clock was associated with an 18% higher risk of death.

*What are the clinical implications?:*

- DNA methylation may complement conventional clinical risk assessment by identifying biological vulnerability beyond established clinical risk factors.
- Multidimensional epigenetic profiling may provide a framework for molecular phenotyping of HF and for identifying patients with residual biological risk who may warrant further investigation.

## Introduction

Heart failure (HF) is a complex clinical syndrome and leading cause of hospitalization, morbidity, and healthcare utilization worldwide.^1^ Although its prevalence rises markedly with age,^2–4^ HF is not simply a consequence of aging, but reflects the cumulative interplay of cardiovascular, inflammatory, metabolic, and systemic processes across the lifespan.^5–8^ Despite improvements in pharmacologic and device-based therapies, HF continues to carry substantial morbidity and mortality, highlighting the need to better understand the biological mechanisms that drive disease progression and identify markers of risk beyond established clinical factors.

Recent advances in high-throughput molecular profiling have enabled the discovery of DNA methylation (DNAm)-based signatures, including epigenetic aging clocks that estimate biological age independently of chronological age.^9–11^ DNAm variation has been associated with lifestyle factors, circulating biomarkers, and mortality, leading to the development of trait-specific DNAm scores.^12–15^ These scores combine information from predefined sets of cytosine-phosphate-guanine (CpG) sites selected in large training datasets to quantify DNAm patterns related to specific biological traits and phenotypes. Unlike circulating biomarkers, which reflect biological states at a single time point, DNAm-based measures capture longer-term exposures and biological processes, potentially providing information beyond contemporaneous biomarker levels. Collectively, these signatures may provide a molecular memory of cumulative exposures and biological aging. Whether they capture clinically relevant prognostic information beyond established risk markers in HF, and whether these associations generalize across diverse global populations, remains unclear.

We therefore evaluated whether a multidimensional panel of 15 established DNAm scores and epigenetic aging measures could characterize biological heterogeneity and provide prognostic information beyond established clinical risk factors and NT-proBNP in a multiethnic cohort of HF patients.

## Methods

### Study design and population

The Global Congestive Heart Failure (G-CHF) registry is a prospective cohort of 23,341 adults (≥18 years) with clinically diagnosed HF, enrolled from 2016 to 2020 across 257 centers in 40 countries. The study design and methodology have been described previously.^16,17^ A molecular substudy included 2,594 participants with baseline whole-blood DNA available for epigenetic profiling. The molecular substudy sample size was determined by the availability of baseline whole-blood DNA samples meeting quality and feasibility requirements within the G-CHF registry; no formal a priori sample-size calculation was performed. Participants were ≥18 years of age and had a clinical diagnosis of HF based on local investigator assessment. Prior evaluations demonstrated that G-CHF represents a well-characterized HF population, based on the prevalence of structural heart disease features, Framingham HF criteria among hospitalized patients, and distribution of NT-proBNP levels.^16^ Baseline data included demographics, HF etiology, New York Heart Association (NYHA) class, comorbidities, smoking, and medications. Ancestry was self-reported and categorized as Caucasian, Latin American, Black African, Bantu/Semi Bantu, Nilotic/Hausa, South Asian, Other Asian, Arab, or Other. Country-income level (low, low-middle, upper-middle, high) was defined at study initiation according to the World Bank country income classification system (2008). The validated MAGGIC risk score was calculated from established clinical variables to estimate mortality risk in patients with HF.^18^ A detailed description of the MAGGIC score and the formula used for calculation is provided in the **Supplementary methods** and **Supplementary Table 1.** The score was analyzed as a continuous variable in statistical models. The study was approved by the local ethics committees, and all participants provided written informed consent.

### Quantification of trait-specific DNAm scores and aging clocks

A custom Illumina Infinium methylation array was used to measure methylation markers corresponding to trait-specific epigenetic scores and aging clocks, as previously described (**Supplementary methods**).^19^ DNAm scores and aging clocks aggregate the methylation level (% of DNA molecules that are methylated at a given site) across multiple trait- and age-associated markers, into a single numerical estimate of an individual’s trait-specific epigenetic signature and biological age. Specifically, trait-specific CpGs for key lifestyle, environmental, inflammatory, and metabolic factors were curated from published DNAm scores. These included scores reflecting lifestyle and environmental exposures (smoking, alcohol intake, alcohol use disorder, Alternative Healthy Eating Index [AHEI], Mediterranean Diet Score [MDS], nitric oxide),^13,20–22^ inflammatory, cardiometabolic, and health span-related traits (C-reactive protein [CRP], body mass index [BMI], frailty, mortality).^14,23–25^ For the AHEI and MDS DNAm scores, higher scores were prespecified to indicate healthier dietary patterns, consistent with the original publication from which the scores were derived.^21^ The directionality of all other DNAm scores was retained as defined in their respective published weight files. DNAm scores for each trait were calculated by extracting trait-specific CpG sites and effect weights from the corresponding weight files and applying these weights to the beta matrix (**Supplementary methods**). The scores were subsequently z-standardized (mean=0, standard deviation [s.d.]=1).

Published aging clocks, including HorvathAge, HannumAge, GrimAge, PhenoAge, and PaceOfAging, were used to estimate biological aging.^9–11,26,27^ Clock values were calculated using established CpG-based algorithms based on published CpG weights and implemented through dedicated analyses pipelines (**Supplementary methods**). Clock-specific epigenetic age acceleration (age gap) was calculated as the residual from regressing biological age on chronological age and subsequently z-standardized to mean=0 and s.d.=1; these standardized values were used in all subsequent analyses.^28^ **Figure 1A** provides an overview of the derivation and standardization of the DNAm scores and aging clocks.

**Figure 1.**
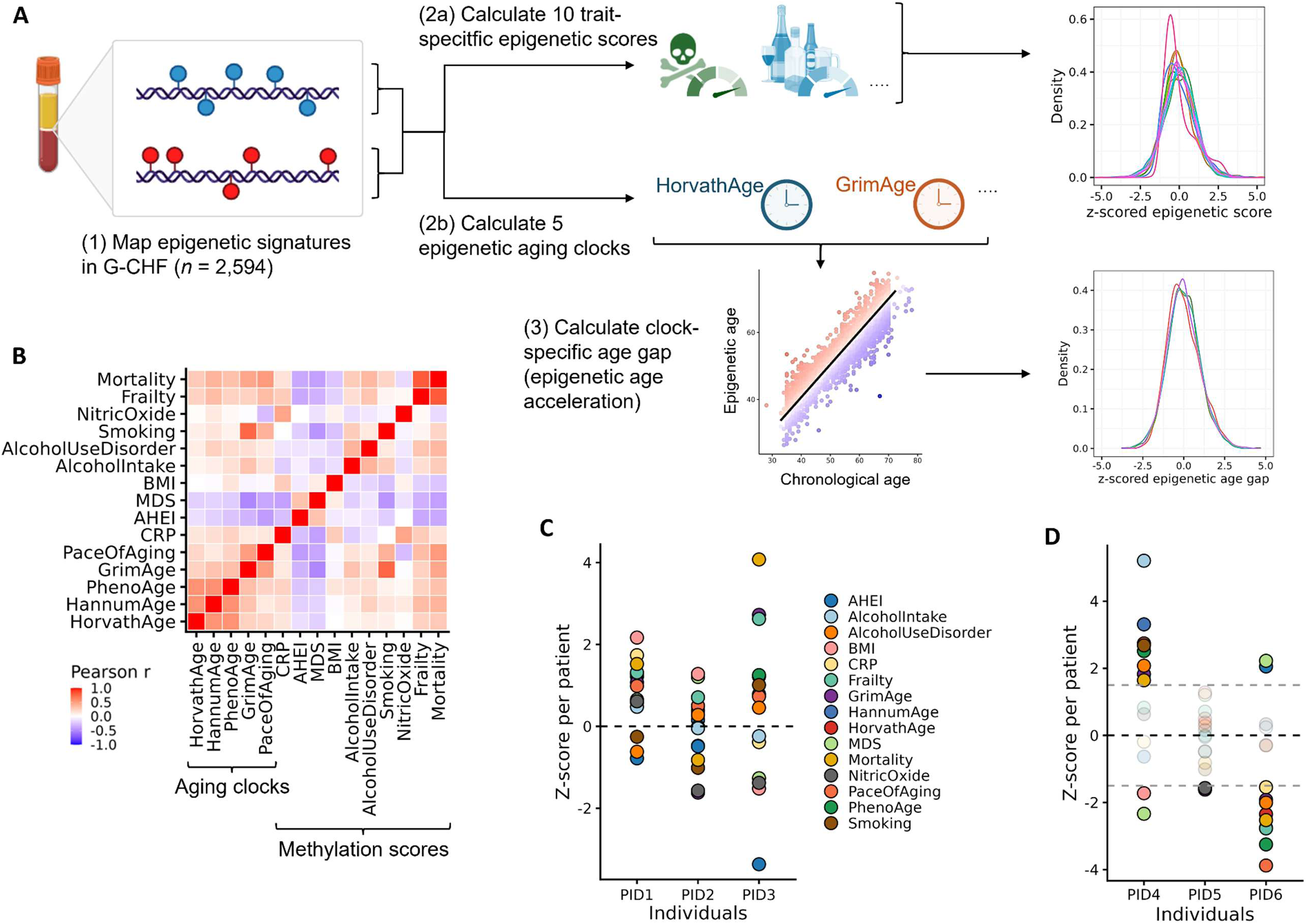
Epigenetic scores and aging clocks in G-CHF. **A**, Study design for deriving trait-specific epigenetic scores and aging clocks from DNAm profiles in the G-CHF registry. Epigenetic scores and aging clocks were calculated in a subset of G-CHF patients with available DNAm data (*n*=2,594). For the aging clocks, epigenetic age acceleration (age gap) was calculated and then z-standardized to allow for direct comparisons with epigenetic scores (also z-score standardized). Epigenetic scores and aging clocks were characterized and tested for associations with lifestyle factors, medications, and clinical outcome risk. **B**, Pairwise correlation of epigenetic scores and aging clocks from all individuals. **C**, Three examples of HF patients (patient ID [PID]) are shown that can have different distribution of epigenetic score/clock profiles. **D**, Three examples of individuals with extreme epigenetic scores/clocks, defined by a 1.5-s.d. increase or decrease in at least one score/clock. Abbreviations: BMI=Body-mass index; MDS=Mediterranean diet score; AHEI=Alternative healthy eating index; CRP=C-reactive protein.

### Clinical outcome assessments

The following outcomes were assessed: HF hospitalization, all-cause mortality, cardiovascular mortality, and a composite of HF hospitalization or all-cause mortality. Patients were followed every 6 months up to 6 years (annual clinic visits and telephone calls at 6 and 18 months), with assessment of vital status and HF hospitalizations at each contact. Deaths were recorded and categorized by local investigators through review of relevant source documents. HF hospitalizations were identified through physician or self-report, with subsequent confirmation by local review of available documentation, and were defined as any HF-related admission that required a hospital stay for at least 24h. Patients were censored at the date of last available follow-up, death, or administrative end of follow-up, as appropriate.

### Statistical analysis

Continuous variables are presented as mean±s.d. and categorical variables as frequencies (percentages). Correlations between z-standardized DNAm scores and aging clocks were evaluated using Pearson’s r correlation coefficients. Associations of DNAm scores and aging clocks with lifestyle and cardiovascular risk factors, as well as HF medication, were assessed using multivariable linear regression. Models for lifestyle and risk factors were adjusted for age, sex, ancestry, and all 8 covariates (education, alcohol intake, hypertension, hyperlipidemia, body mass index, diabetes, smoking, waist-to-hip ratio). Models for HF medication use were adjusted for age, sex, ancestry, and all 9 medication classes (loop diuretics, digoxin, mineralocorticoid receptor antagonist [MRA], betablocker, antiarrhythmics, calcium channel blockers, sacubitril/valsartan, angiotensin-converting enzyme [ACE] inhibitors, and angiotensin II receptor blockers [ARBs]). β coefficients and two-sided P values were estimated for each association, with adjustment for multiple testing using the Benjamini-Hochberg method. Associations of smoking and alcohol intake DNAm scores with their corresponding behavioral phenotypes were assessed using linear trend tests across ordered categories.

Associations of z-standardized DNAm scores and aging clocks with clinical outcomes were evaluated per 1-s.d. increase using Cox proportional hazards models adjusted for age, sex, ancestry, the MAGGIC score, and NT-proBNP. The proportional hazards assumption was assessed with Schoenfeld residuals, which indicated no violations. Similar analyses were performed according to clinical risk, defined by the population median MAGGIC score (low risk, MAGGIC <17; high risk, MAGGIC ≥17), and by HF phenotype, categorized as HF with preserved ejection fraction (HFpEF), HF with mildly reduced ejection fraction (HFmrEF), and HF with reduced ejection fraction (HFrEF). Differences in associations between MAGGIC groups were assessed using DNAm score/clock-by-MAGGIC group interaction terms in Cox models adjusted for age, sex, ancestry, and NT-proBNP. Patients were categorized into quartiles of the Mortality DNAm score. The highest quartile (Q4; ≥75th percentile) was compared with the lower 3 quartiles (Q1-Q3; <75th percentile) using Kaplan-Meier curves and multivariable Cox models adjusted for the same covariates as described above. Analyses were performed in the overall cohort and repeated in patients with low clinical risk (MAGGIC <17).

Extreme values of all DNAm scores and aging clocks were examined in relation to all-cause mortality. Extreme values were defined as standardized values >1.5 or <-1.5 s.d. and classified according to their direction of association with mortality. For scores for which higher values indicated greater risk, >1.5 s.d. was considered a higher-risk extreme and <-1.5 s.d. a lower-risk extreme; for protective scores, this direction was reversed. Higher- and lower-risk extreme counts were then calculated across all 15 measures and patients were subsequently categorized according to the number of extreme measures. The association between accumulation of higher- or lower-risk extreme DNAm scores or aging clocks and all-cause mortality was evaluated using multivariable-adjusted Cox models and Kaplan-Meier curves. All analyses were conducted using complete cases for the variables included in each model. All statistical tests were 2-sided, and a P value <0.05 was considered statistically significant. Statistical analyses were performed using R version 4.4 (R Foundation for Statistical Computing, Vienna, Austria).

## Results

Baseline characteristics of the study population are presented in **Table 1**. Mean age was 62.7±14.0 years, 66.2% were male, mean left ventricular ejection fraction was 40.1±14.1%, and participants had overall a high burden of cardiovascular comorbidities. Pairwise correlation analyses revealed moderate positive correlations between epigenetic aging clocks and DNAm-based mortality and frailty scores (*r*=0.23-0.52), whereas diet-related scores (MDS and AHEI) were inversely correlated with most other scores and clocks (**Figure 1B**). At the individual level, substantial heterogeneity was observed across DNAm scores and aging clocks, with several individuals exhibiting marked discordance between domains (**Figure 1C**). For example, some participants displayed elevated aging clock estimates and inflammation- or mortality scores alongside favorable diet-related scores, whereas others showed the opposite pattern.

**Table 1.** Baseline characteristics of study population.

| Characteristic | N=2594 |
| --- | --- |
| Age (years), mean±SD | 62.7±14.0 |
| Male sex, n (%) | 1716 (66.2) |
| Ancestry, n (%) |  |
| Caucasian | 1435 (55.3) |
| Latin American | 405 (15.6) |
| Black African | 277 (10.7) |
| Bantu/Semi Bantu | 152 (5.9) |
| Nilotic/Hausa | 120 (4.6) |
| Other Asian | 49 (1.9) |
| South Asian | 7 (0.3) |
| Arab | 15 (0.6) |
| Other | 134 (5.2) |
| Etiology of HF, n (%) |  |
| Ischemic | 969 (37.4) |
| Hypertensive | 509 (19.6) |
| Idiopathic | 407 (15.7) |
| Valvular | 98 (3.8) |
| Other | 611 (23.6) |
| Smoking, n (%) |  |
| Never | 1337 (51.5) |
| Former | 1019 (39.3) |
| Current | 238 (9.2) |
| Alcohol use, n (%) |  |
| Never | 1134 (43.7) |
| Former | 587 (22.6) |
| Current | 873 (33.7) |
| NYHA class, n (%) |  |
| I | 407 (15.7) |
| II | 1519 (58.8) |
| III | 572 (22.1) |
| IV | 87 (3.4) |
| LV ejection fraction (%), mean±SD | 40.1±14.1 |
| SBP (mmHg), mean±SD | 123±19 |
| DBP (mmHg), mean±SD | 75±12 |
| BMI (kg/m<sup>2</sup>), mean±SD | 28.7±6.0 |
| Serum creatinine (μmol/L), mean±SD | 107.3±67.6 |
| Comorbidities, n (%) |  |
| Hypertension | 1856 (71.6) |
| Diabetes | 722 (27.8) |
| CAD | 1037 (40.0)403 |
| Atrial fibrillation | 765 (29.5) |
| Chronic obstructive pulmonary disease | 266 (10.3) |
| MAGGIC risk score, mean±SD | 16.7±7.0 |
| Taking ACE inhibitors/ARB, n (%) | 1957 (75.5) |
| Taking β-blockers, n (%) | 2195 (84.7) |
| Taking mineralocorticoid receptor antagonist, n (%) | 1587 (61.2) |
| Abbreviations: ARB=angiotensin receptor blocker, CAD=coronary artery disease, DBP=diastolic blood pressure, LV=left ventricular, SBP=systolic blood pressure.<br>Missing data: NYHA class (n=9). |  |

In multivariable linear models, several scores were associated with cardiometabolic and behavioral factors, including smoking, alcohol intake, waist-to-hip ratio, hypertension, hyperlipidemia, diabetes, and education (**Figure 2A**). Mortality-, frailty-, and aging-related epigenetic scores/clocks showed strong associations with smoking and waist-to-hip ratio (β=-0.21-0.98), whereas favorable diet-related scores were inversely associated with cardiovascular risk factors. In exploratory analysis, we tested the associations between all fifteen epigenetic scores and clocks and nine HF-specific medications (**Figure 2B**). Loop diuretics and digoxin use showed the most pronounced associations with accelerated biological aging and were inversely associated with healthy dietary indices (MDS and AHEI, ranging from β=-0.131 to -0.215). Conversely, Sacubitril/Valsartan (Sac/Val) use was associated with a distinct epigenetic signature, most notably a negative association with CRP and mortality scores (β=-0.201 and - 0.127). A stepwise increase in smoking-related DNAm scores was observed across never, former, and current smokers (P_trend_<0.001) (**Figure 2C**, **Supplementary Figure 1**). Similarly, the alcohol intake DNAm score was higher among current drinkers compared to former and never drinkers (P_trend_<0.001) (**Figure 2D, Supplementary Figure 2**). ARB use was associated with lower epigenetic age across all aging clocks except HorvathAge, compared with no ARB use (**Figures 2B and 2E**).

**Figure 2.**
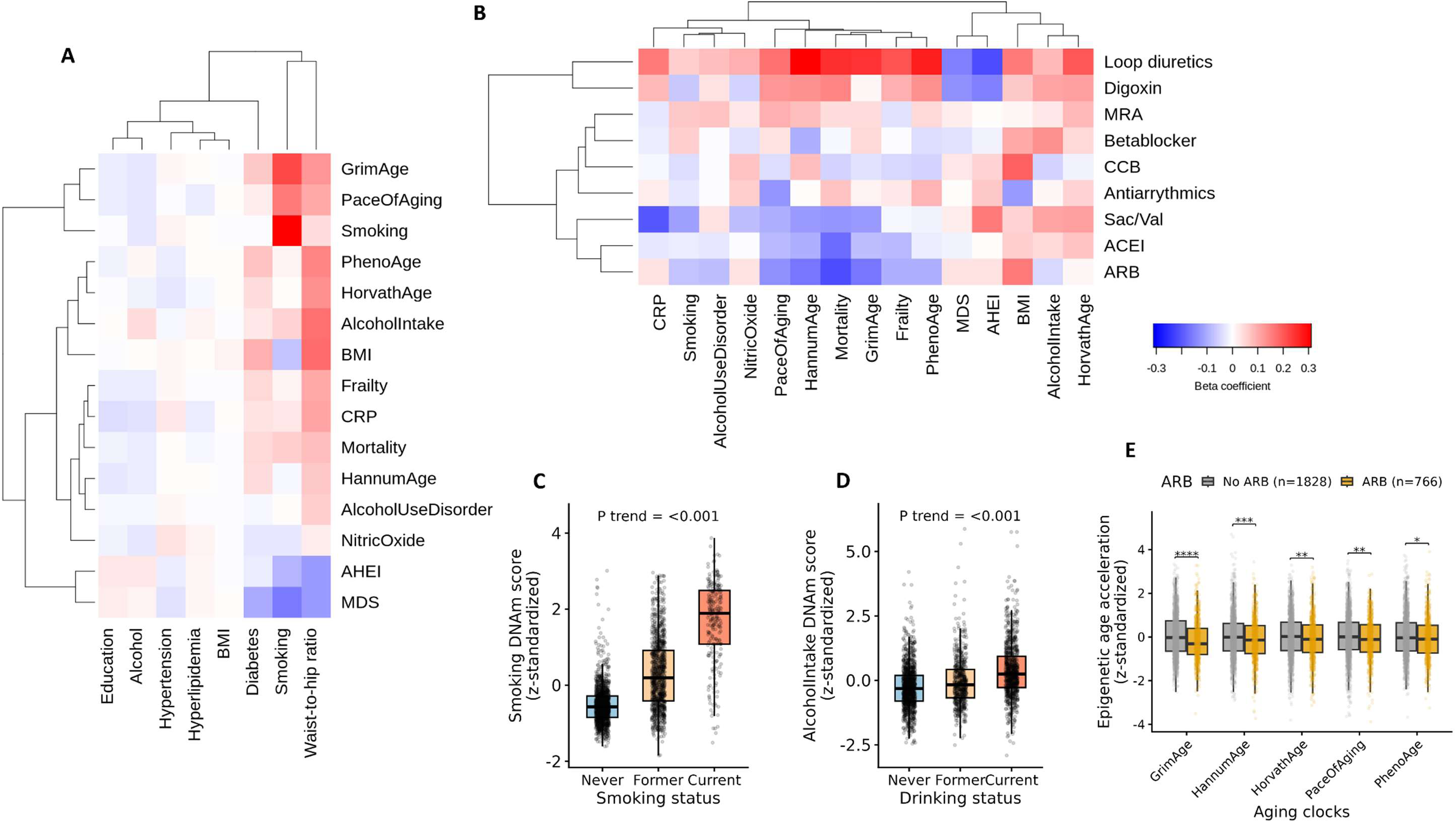
Associations of epigenetic scores and aging clocks with lifestyle factors and medications. **A**, Multivariable linear regression was used to determine the association between epigenetic scores and aging clocks and modifiable lifestyle and cardiovascular risk factors. Models were adjusted for age, sex, ancestry, and all 8 factors. The heatmap is color coded by the beta coefficient. **B**, Multivariable linear regression was used to determine the association between epigenetic scores and aging clocks and medication intake. Models were adjusted for age, sex, ancestry, and all 9 medication classes. In both analyses, P values were adjusted using the Benjamini-Hochberg method and non-significant associations were displayed in white. **C**,**D**, Box plots showing z-standardized DNAm scores for smoking and alcohol intake, stratified by baseline smoking and alcohol consumption status (never, former, current). Boxes represent the interquartile range (IQR), center lines denote the median, whiskers extend to 1.5×IQR, and points indicate individual participants. **E**, Box plots of epigenetic aging clocks in individuals stratified by ARB treatment. P values are derived from two-sided t-tests comparing groups. Abbreviations: MRA=Mineralocorticoid receptor antagonists; CCB=Calcium channel blocker; Sac/Val=Sacubitril/Valsartan; ACEI=Angiotensin-converting enzyme (ACE) inhibitors; ARB=Angiotensin II receptor blockers; ****= P<0.0001; ***=P<0.001; **=P<0.01; *=P<0.05.

During a median follow-up of 3.0 years (interquartile range, 1.7-3.6), 338 patients (13%) were hospitalized for HF, 349 (13%) died from cardiovascular causes, and 565 (22%) died from any cause. Across all clinical outcomes, higher epigenetic age, inflammation, and frailty were generally associated with increased risk, whereas higher diet epigenetic scores were associated with lower risk (**Figure 3**). The strongest associations were observed for GrimAge, CRP, Frailty, and the Mortality score. For all-cause death, adjusted hazard ratios (aHR) per 1-s.d. increase were 1.36 (95% CI, 1.24-1.50) for GrimAge, 1.27 (95% CI, 1.17-1.38) for CRP, 1.44 (95% CI, 1.28-1.63) for Frailty, and 1.48 (95% CI, 1.32-1.65) for the Mortality score. Similar associations were observed for cardiovascular death, with aHRs of 1.38 (95% CI, 1.22-1.56), 1.28 (95% CI, 1.15-1.42), 1.55 (95% CI, 1.32-1.80), and 1.52 (95% CI, 1.32-1.76), respectively. In contrast, higher AHEI and MDS scores were consistently associated with lower risk across outcomes, including all-cause death (aHR, 0.81 [95% CI, 0.75-0.88] and 0.86 [95% CI, 0.79-0.94]) and cardiovascular death (aHR, 0.78 [95% CI, 0.70-0.86] and 0.82 [95% CI, 0.73-0.91]). Associations for HF hospitalization and the composite of HF hospitalization or death were directionally consistent but generally weaker. When stratified by MAGGIC score, some associations between DNAm scores/aging clocks and outcomes were stronger among patients with lower clinical risk (MAGGIC <17; **Supplementary Figure 3**). This pattern was most apparent for mortality outcomes, particularly for Frailty, Mortality, and CRP DNAm scores, for which HRs were consistently higher among patients with lower MAGGIC scores (P_interaction_<0.05). Associations with HF hospitalization and the composite of HF hospitalization or death showed a similar pattern, although less pronounced. Baseline LVEF was available in 2,174 of 2,594 patients. Among these patients, a similar pattern of associations across HF types was observed, although effect sizes were generally smaller in HFrEF patients (**Supplementary Figure 4**).

**Figure 3.**
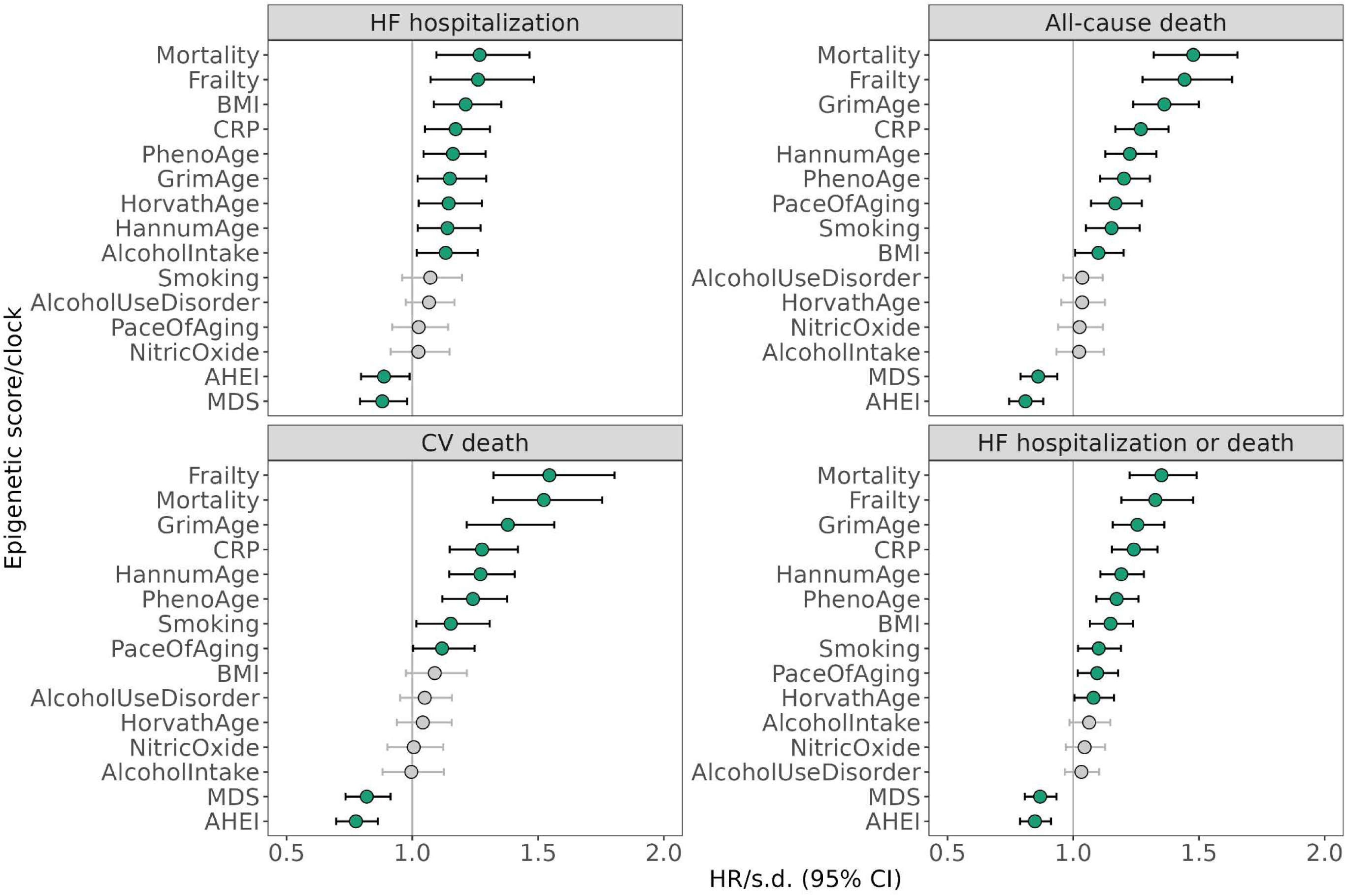
Associations of epigenetic scores and aging clocks with clinical outcomes. Multivariable Cox proportional hazards models were fitted for each z-standardized epigenetic score and aging clock to estimate associations with HF hospitalization, all-cause death, cardiovascular death, and composite of HF hospitalization or all-cause death, adjusting for age, sex, ancestry, the MAGGIC score, and NT-proBNP. Hazard ratios are reported per 1-s.d. increase.

The Mortality DNAm score showed the strongest and most consistent associations with clinical outcomes and was therefore selected for downstream analyses. Patients in the highest quartile of the Mortality DNAm score (Q4) had a higher estimated probability of experiencing each clinical outcome compared with those in the lower quartiles (Q1-Q3; **Figure 4**). Similar associations were observed among patients with lower clinical risk (MAGGIC <17). In multivariable analyses, Q4 was associated with a higher risk of HF hospitalization (aHR, 1.54 [95% CI, 1.19-2.00]), all-cause death (aHR, 1.73 [95% CI, 1.41-2.13]), CV death (aHR, 2.10 [95% CI, 1.59-2.76]), and HF hospitalization or death (aHR, 1.50 [95% CI, 1.26-1.79]) compared with Q1-Q3 (**Supplementary Table 2**). The associations were more pronounced among patients with MAGGIC <17, with aHRs of 1.59 (95% CI, 0.98-2.58) for HF hospitalization, 2.70 (95% CI, 1.84-3.96) for all-cause death, 4.22 (95% CI, 2.59-6.86) for CV death, and 2.00 (95% CI, 1.44-2.76) for HF hospitalization or death.

**Figure 4.**
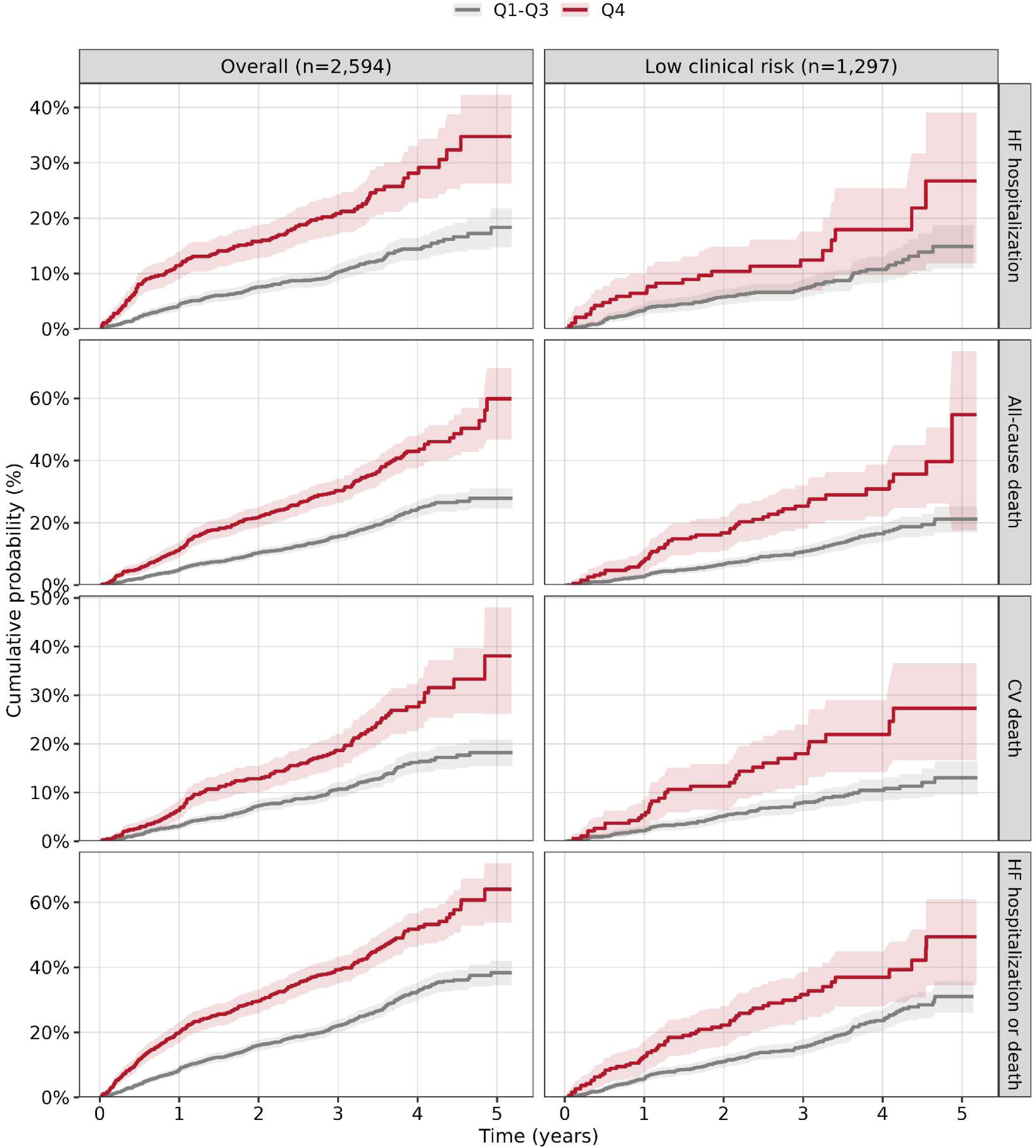
Mortality DNAm score and cumulative probability of clinical outcomes in the overall cohort and in patients with low clinical risk. Kaplan-Meier estimates of the cumulative probability of HF hospitalization, all-cause death, cardiovascular death, and HF hospitalization or death according to Mortality DNAm score quartiles (Q1-Q3 versus Q4). Estimates are shown for the overall cohort (n=2,594; left) and among patients with low clinical risk (MAGGIC score <17; n=1,297; right). Shaded areas represent 95% confidence intervals.

Next, we identified patients with extreme DNAm profiles, defined as scores or clocks exceeding ±1.5 s.d. from the population mean in any domain (**Supplementary Figure 5**). Overall, 54% exhibited at least one extreme-high and 46% exhibited at least one extreme-low signature. A graded relationship was observed, with progressively higher mortality risk among patients with increasing numbers of extreme high values (**Figure 5A**). Patients with 4-5 high extremes showed the greatest risk (aHR 2.27, 95% CI 1.65-3.13) compared with those without extreme scores. In contrast, higher number of extreme low scores/clocks was not associated with mortality risk. Cumulative survival curves across number of extreme scores are shown in **Figure 5B**. Each additional extreme epigenetic score/aging clock was associated with an 18% higher risk of death (aHR, 1.18; P=1.82×10⁻⁹).

**Figure 5.**
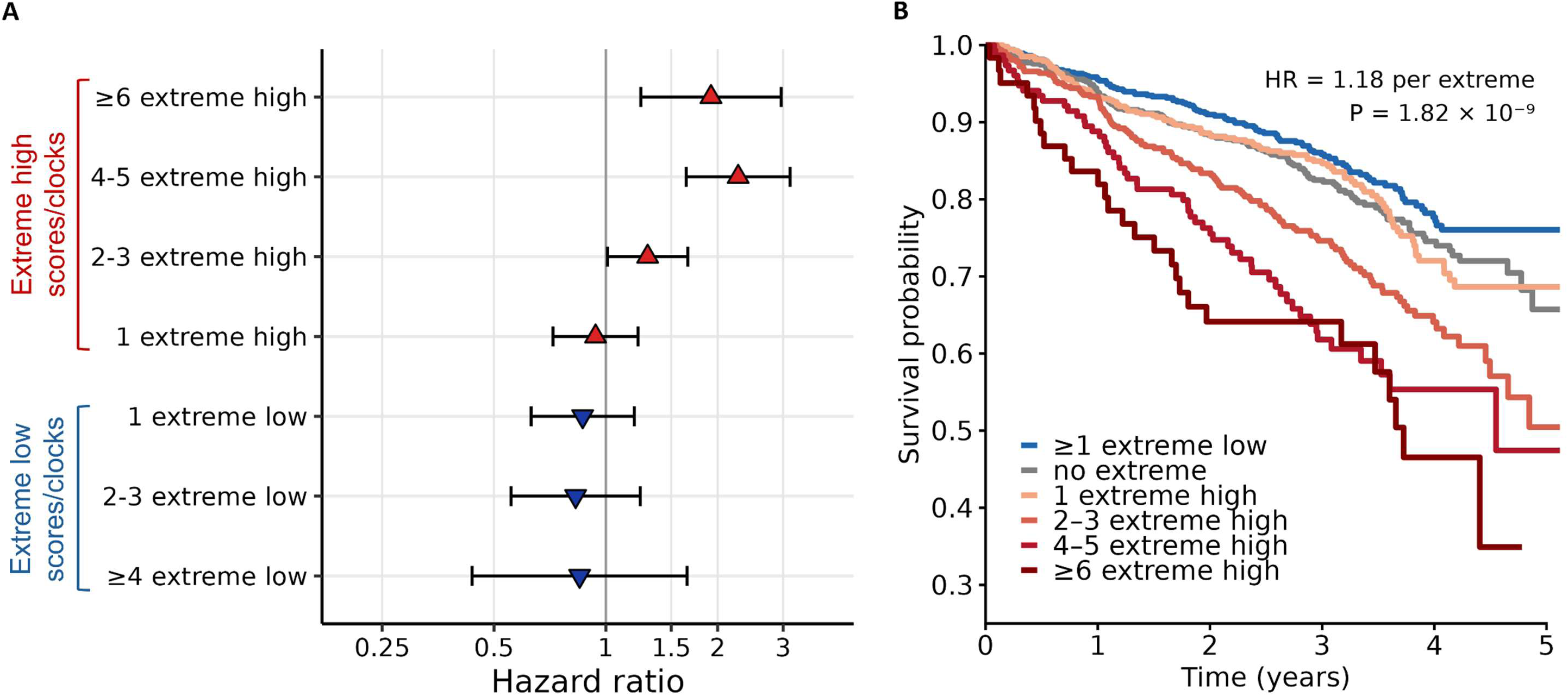
Cumulative burden of epigenetic scores and aging clocks and mortality risk A, Multivariable Cox proportional hazards models were fitted to assess the association of the number of extreme high (>1.5 s.d.) and extreme low (<-1.5 s.d.) epigenetic scores and aging clocks with all-cause death, adjusted for age, sex, ancestry, the MAGGIC score, and NT-proBNP. **B**, Kaplan-Meier survival curves according to the number of extreme high and low epigenetic score and aging clocks. Patients were categorized according to the number of epigenetic scores/clocks exceeding 1.5 s.d. The hazard ratio represents the risk of death per one additional extreme measure, estimated from a multivariable Cox model adjusted for age, sex, ancestry, MAGGIC score, and NT-proBNP.

## Discussion

In this multinational cohort of HF patients, DNAm profiling identified biological heterogeneity not fully represented by conventional clinical risk assessment. DNAm scores and aging clocks were associated with lifestyle exposures, HF medication use, and clinical outcomes, with several measures providing prognostic information after adjusting for the MAGGIC risk score and NT-proBNP. These associations were more pronounced among patients at lower clinical risk, and a greater cumulative burden of adverse epigenetic measures was associated with substantially higher mortality. Importantly, these associations persisted after adjustment for chronological age, indicating that the aging-related signatures captured biological variation beyond chronological age. Together, these findings suggest that DNAm profiling may provide complementary information on biological vulnerability and residual risk in HF.

The association of DNAm scores with modifiable lifestyle exposures supports their biological relevance in HF. We found markedly higher smoking DNAm scores among current smokers, whereas former smokers had intermediate scores between current and never smokers. These findings are consistent with prior studies showing that smoking-associated DNAm signatures are dynamic and can partially normalize following smoking cessation.^15,29,30^ The pattern observed in former smokers suggests that DNAm signatures may reflect not only cumulative exposure but also changes associated with smoking cessation and time since exposure. A similar pattern was observed for alcohol intake, consistent with previous reports of reversible DNAm alterations after alcohol withdrawal.^31,32^ These findings highlight the sensitivity of DNAm-based scores to lifestyle behaviors and raise the possibility that they could serve as molecular markers of exposure history.

We observed significant associations between HF medication use and epigenetic aging. ARB and ACE-I, and to a lesser extent sacubitril/valsartan use, were associated with lower measures of biological aging across multiple clocks. These findings are consistent with experimental and observational evidence linking renin-angiotensin system modulation to pathways involved in biological aging.^33–35^ However, these analyses were observational and may be affected by confounding by indication, disease severity, and other differences between patients receiving therapies. The observed associations should therefore not be interpreted as evidence that neurohormonal blockade reverses biological aging. These findings are hypothesis-generating and warrant evaluation in longitudinal studies examining whether HF therapies influence longitudinal trajectories of biological aging.

A central finding of this study was that multiple DNAm scores and aging clocks were associated with HF hospitalization and mortality after adjustment for both the MAGGIC risk score and NT-proBNP. Epigenetic measures related to health span, particularly the Mortality and Frailty DNAm scores, together with inflammatory measures and aging clocks, showed the strongest associations. These findings are consistent with prior studies identifying GrimAge and CRP-related DNAm score as robust predictors of mortality.^36,37^ Importantly, the associations were more pronounced among patients with lower MAGGIC scores. This suggests that epigenetic measures may be informative in patients who are deemed lower risk based on conventional risk factors but may have substantial underlying biological vulnerability. Although these findings require confirmation, they raise the possibility that molecular profiling could complement, rather than replace, established clinical risk assessment by identifying residual risk not fully captured by conventional clinical measures.

The association between the cumulative burden of extreme epigenetic measures and mortality provides support for the concept that multiple dimensions of biological vulnerability may converge in HF. Rather than reflecting a single pathway, the 15 DNAm scores and aging clocks capture several domains, including smoking and alcohol exposure, diet, inflammation, adiposity, frailty, mortality risk, and biological aging. Patients with multiple extreme measures had substantially higher mortality risk than those without extreme measures, suggesting that adverse signals across several biological domains may identify a vulnerable HF phenotype. This cumulative approach may therefore be more informative than considering individual epigenetic measures in isolation. Together, these findings support the potential value of integrating complementary epigenetic measures to characterize residual risk in HF. However, whether such measures improve clinical decision-making or patient outcomes remains to be investigated in future studies.

This study has some limitations. First, DNAm profiles were measured at a single baseline time point, precluding assessment of longitudinal changes and limiting causal inference regarding temporal relationships between epigenetic signatures, exposures, and mortality risk. Second, although the cohort was large and multinational, participants in the molecular substudy were selected based on biospecimen availability, which may introduce selection bias. Third, DNAm was quantified in whole blood, which may not fully reflect tissue-specific processes relevant to cardiac remodeling and HF progression. Fourth, associations with lifestyle factors and medication use were observational and susceptible to residual confounding. Finally, although associations were evaluated across clinical risk strata and HF types in this multinational and multiethnic population, the generalizability of the observed associations and incremental discriminatory value of these established DNAm measures requires confirmation in independent HF cohorts.

In summary, DNAm-based scores and aging clocks capture multiple dimensions of biological heterogeneity in HF and provide prognostic information beyond clinical risk factors and NT-proBNP. Future studies should determine whether longitudinal changes in these signatures reflect treatment response or changes in modifiable exposures, and whether incorporating molecular phenotyping into clinical care can improve risk stratification and inform management in HF.

## Data Availability

Individual-level data are not available to the public as per the study protocol and data-sharing policies. Summary data and the statistical code can be made available on reasonable request.

## Ethical Approval

The study was approved by the local ethics committees, and all participants provided written informed consent.

## Acknowledgments

The authors gratefully acknowledge the invaluable contributions of the G-CHF investigators. A full list of the investigators is provided in the Supplement.

